# Estimating post-treatment recurrence after multidrug-resistant tuberculosis treatment among patients with and without HIV: the impact of assumptions about death and missing follow-up

**DOI:** 10.1101/2023.05.24.23290472

**Authors:** Sara M. Sauer, Carole D. Mitnick, Uzma Khan, Catherine Hewison, Mathieu Bastard, David Holtzman, Stephanie Law, Munira Khan, Shrivani Padayachee, Saman Ahmed, Afshan K. Isani, Aga Krisnanda, Stalz Charles Vilbrun, Sagit Bektasov, Andargachew Kumsa, Wisney Docteur, Karen Tintaya, Mark McNicol, Hakob Atshemyan, Tatiana Voynilo, Thin Thin Thwe, Kwonjune Seung, Michael Rich, Helena Huerga, Palwasha Khan, Molly Franke

## Abstract

**Background:** Quantification of recurrence risk following successful treatment is crucial to evaluating regimens for multidrug- or rifampicin-resistant (MDR/RR) tuberculosis (TB). However, such analyses are complicated when some patients die or become lost during post-treatment-follow-up.

**Methods:** We analyzed data on 1,991 patients who successfully completed a longer MDR/RR-TB regimen containing bedaquiline and/or delamanid between 2015 and 2018 in 16 countries. Using five approaches for handling post-treatment deaths, we estimated the six-month post-treatment TB recurrence risk overall, and by HIV status. We used inverse-probability-weighting to account for patients with missing follow-up and investigated the impact of potential bias from excluding these patients without applying inverse-probability weights.

**Results:** The estimated TB recurrence risk was 6.6 per 1000 (95% confidence interval (CI):3.2,11.2) when deaths were handled as non-recurrences, and 6.7 per 1000 (95% CI:2.8,12.2) when deaths were censored and inverse-probability weights were applied to account for the excluded deaths. The estimated risk of composite recurrence outcomes were 24.2 (95% CI:14.1,37.0), 10.5 (95% CI:5.6,16.6), and 7.8 (95% CI:3.9,13.2) per 1000 for recurrence or 1) any death, 2) death with unknown or TB-related cause, 3) TB-related death, respectively. Corresponding relative risks for HIV status varied in direction and magnitude. Exclusion of patients with missing follow-up without inverse-probability-weighting had a small but apparent impact on estimates.

**Conclusion:** The estimated six-month TB recurrence risk was low, and the association with HIV status was inconclusive due to few recurrence events. Estimation of post-treatment recurrence will be enhanced by explicit assumptions about deaths and appropriate adjustment for missing follow-up data.

## BACKGROUND

Quantifying post-treatment tuberculosis (TB) recurrence is critical to evaluating the effectiveness of all multidrug- and rifampicin-resistant (MDR/RR) TB regimens during routine care. Estimating risk of recurrence is complicated, however, when some patients die or are missing data during the post-treatment follow-up (FU) period. Post-treatment deaths that are unrelated to TB preclude recurrence and constitute *competing* [1-3] or *intercurrent* [4] events. Conversely, if deaths occur due to undetected TB, the risk of recurrence can be underestimated. In analyses of recurrence, assumptions are made regarding post-treatment deaths; different assumptions alter the estimated quantity and the resulting interpretation [1]. Prior studies have typically 1) excluded patients who died [5-8], which will bias the estimated risk of recurrence in the likely scenario that people who do and do not die have different underlying risks of TB recurrence; 2) effectively assumed that none of the post-treatment deaths were due to recurrent TB [9-10], which is not supported by the literature [11-13]; or 3) omitted details on how deaths were analyzed [14-18], which prevents a clear interpretation of the resulting estimate.

Analytic challenges also arise from patients for whom there is no post-treatment follow-up data. In these patients, it is unknown whether recurrent TB occurred. Most prior studies that estimate TB recurrence have excluded individuals who lack post-treatment follow-up [5-7]. Valid estimates under this approach require that individuals who do and do not have post-treatment follow-up have a similar underlying risk of recurrent TB. If, on the other hand, there are common causes of recurrence and lack of post-treatment follow-up, this assumption does not hold, and simply excluding individuals who lack follow-up data may lead to selection bias [19-20].

In this paper we estimated the overall risk of TB recurrence within six months of successful MDR/RR-TB treatment with a longer regimen containing bedaquiline (BDQ) and/or delamanid (DLM) and compared this risk among those with and without HIV infection. Compared to people without HIV, those with HIV are more susceptible to TB disease [21], at increased risk of MDR-TB [22], more prone to serious adverse events [23], and at higher risk of unfavorable treatment outcomes, including recurrence [24-25]. In our analyses, we examined multiple approaches for handling post-treatment deaths or adjustment for potential bias due to exclusion of missing follow-up data and assessed whether results were sensitive to the approach chosen.

## METHODS

### Data source and study population

We used data from the endTB Observational Study, a prospective cohort of 2,788 individuals who initiated a longer individualized MDR/RR-TB regimen containing BDQ and/or DLM between 2015 and 2018 in one of seventeen countries (Armenia, Bangladesh, Belarus, Ethiopia, Georgia, Haiti, Indonesia, Kazakhstan, Kenya, Kyrgyzstan, Lesotho, Myanmar, North Korea, Pakistan, Peru, South Africa, Vietnam) on five continents [26]. For this analysis, we included patients’ first treatment in endTB, excluded those with confirmed sensitivity to rifampicin and restricted to patients with end of treatment success, defined as cure or treatment completion. We additionally excluded all individuals from North Korea, as post-treatment follow-up was not conducted at this site.

### Outcome measure

The outcome of interest was a binary indicator of TB recurrence, irrespective of whether it was reinfection with a new strain of *Mycobacterium tuberculosis* or relapse with the initial infecting strain, within six months of successful treatment. Across participating sites, post-treatment outcomes and culture results were recorded by clinicians on a standardized form at scheduled visits six months post-treatment. These post-treatment outcomes were classified into the following categories: no change, post-treatment death, recurrent TB, lost to follow-up (LTFU), and not evaluated. We grouped individuals with the latter two outcomes, or for whom there was no post-treatment outcome form, into a single ‘missing FU’ category.

### Statistical methods

#### Addressing post-treatment deaths

We considered five strategies from the competing/intercurrent events literature for handling deaths that occurred within six months post-treatment (Table 1, Figure 1).

**Table 1:** Strategies for handling deaths post-treatment

| Approach | Operationalization | Assumptions |
| --- | --- | --- |
| **Observed recurrence**<sup>1</sup> | Include individuals who died in the analysis and treat as non-recurrences<sup>4</sup> | Patients who died did *not* experience recurrent TB |
| **Hypothetical recurrence**<sup>2</sup> | Censor (exclude) individuals who died, account for common causes of death and recurrence with IP weights | All common causes of death and recurrence have been accounted for |
| **Observed composite/any**<sup>3</sup> | Define a composite outcome of recurrence or death from *any* cause |  |
| **Observed composite/probable**<sup>3</sup> | Define a composite outcome of recurrence or death with either 1) unknown cause, 2) cause related to TB; treat deaths with cause unrelated to TB as non-recurrences<sup>4</sup> | Patients with deaths unrelated to TB did *not* experience recurrent TB |
| **Observed composite/explicit**<sup>3</sup> | Define a composite outcome of recurrence or death with cause related to TB, treat deaths with 1) unknown cause and 2) cause unrelated to TB as non-recurrences<sup>4</sup> | Patients with deaths of unknown cause or cause unrelated to TB did *not* experience recurrent TB |

<sup>1</sup>when estimating the association between HIV and post-treatment recurrence, the *observed recurrence* strategy corresponds to estimating the *total effect* of HIV in the causal inference with competing events literature [1-3]
<sup>2</sup>in the causal inference literature, the quantity estimated through the hypothetical recurrence strategy is known as the *controlled direct effect* [1-3]; in the intercurrent events literature, the corresponding strategy is called the *hypothetical* strategy [4]
<sup>3</sup>composite strategies correspond to estimating the *total effect on a composite event* in the causal inference literature [1-3], and as the *composite* strategy in the corresponding intercurrent events literature [4]
<sup>4</sup>recurrent TB outcome indicator set to 0 for all patients handled as non-recurrences

**Figure 1:**
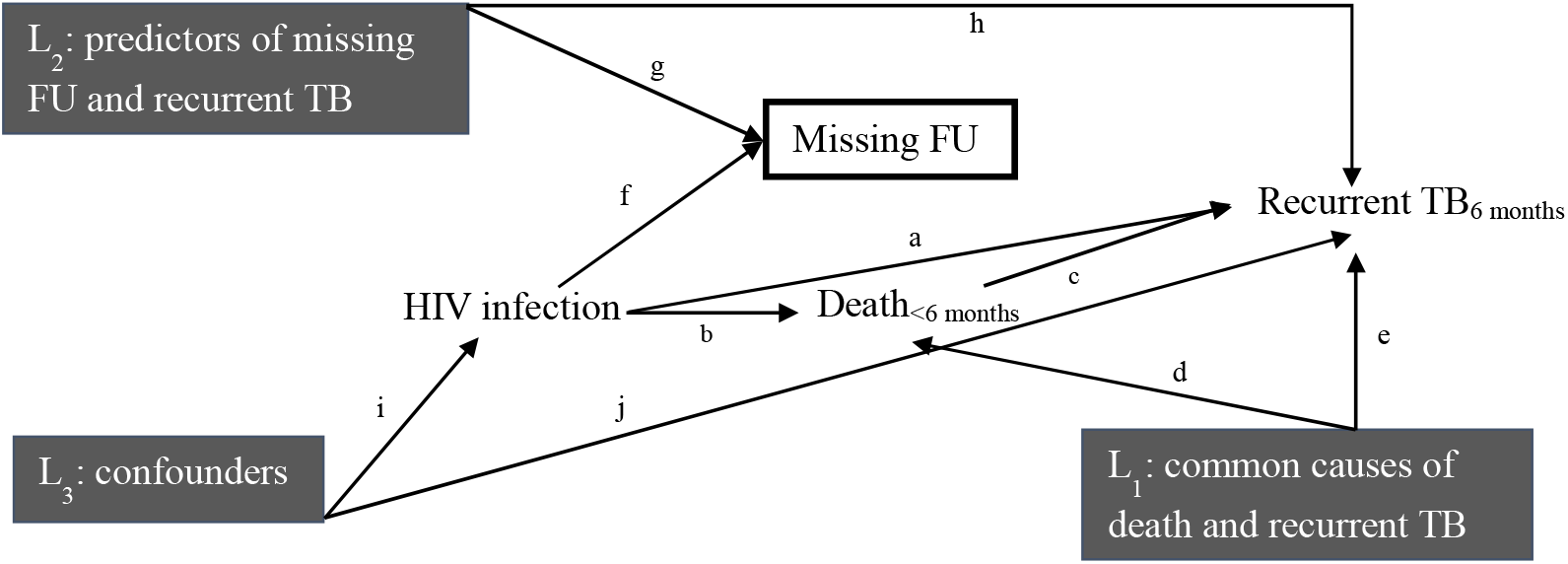
Graphical representation of the association between HIV and recurrent TB at six months post-treatment. The *observed recurrence* strategy estimates paths *a* and *b-c*; the *hypothetical recurrence* strategy estimates path *a* only, adjusting for common causes of death within six months post-treatment (prior to recurrence) and recurrent TB (L_1_) to eliminate bias through the *b-d-e* path; adjustment for the joint predictors of missing FU (L_2_) accounts for potential selection bias through the *f-g-h* path; adjustment for common causes of HIV infection and recurrent TB (L_3_) addresses potential confounding bias by blocking the *i-j* path.TB

First, in what we term the *observed recurrence* strategy, we included patients who died but did not count them as recurrences, thereby assuming that patients who died did *not* experience recurrent TB. Second, in the *hypothetical recurrence* strategy [1-4], we excluded patients who died and applied stabilized inverse-probability (IP) weights to account for potentially different underlying risks of recurrent TB among excluded vs included patients. This strategy posits a hypothetical scenario in which all deaths were prevented, so that whether these (in reality deceased) patients experienced recurrent TB within six months was observed. Factors associated with both post-treatment death and post-treatment recurrence were used in the IP weight construction, namely age, CD4 count, cavitary disease, alcohol use, drug use, smoking, incarceration, diabetes, stoppage of group A drugs (BDQ, fluoroquinolones, linezolid) due to adverse events, and BMI (see Supporting Information for details on weight construction).

In strategies 3 through 5, we defined various composite outcomes of recurrence or death. In the *observed composite/any* strategy, we included recurrent TB and death from any cause in the outcome, while in the *observed composite/probable* strategy, we included recurrent TB and death with unknown or TB-related cause. Finally, in the *observed composite/explicit* strategy, we included recurrent TB and only death due to a TB-related cause in the outcome. In the latter two composite outcome strategies, we included any deaths not counted in the outcome as non-recurrences, thereby assuming that these patients did not experience recurrent TB.

#### Addressing missing post-treatment follow-up data

We utilized stabilized IP weights to overcome the potential selection bias induced by excluding patients who lack follow-up data [19-20]. We constructed the weight denominators by estimating the probabilities of having an observed six month post-treatment outcome conditional on HIV status and joint predictors of missing FU and recurrent TB: cavitary disease, BMI, time to culture conversion, use and duration of Group A drugs, stoppage of Group A drugs due to an adverse event, likely effectiveness of Group A drugs, country of treatment site, and diabetes. See Supporting Information for details.

#### Estimating post-treatment recurrence

We computed estimates of recurrent TB risk, overall and by HIV status, by fitting an IP-weighted logistic regression model with the HIV status indicator as the only covariate. Weights to account for missing FU were included in all analyses, whereas weights to account for common casues of death and recurrent TB were only employed in the *hypothetical recurrence* analysis, in which new overall weights were constructed by multiplying together the sets of weights. Furthermore, we incorporated an additional set of IP weights in all analyses to adjust for potential confounding by the common causes of HIV status and recurrent TB, in this case age, country of treatment site, hepatitis C, alcohol use, drug use, smoking, and incarceration (see Supporting Information). We constructed 95% credible intervals for all estimates with a bootstrapping approach [27] using the `boot` package in R.

#### Sensitivity analysis

We conducted a sensitivity analysis to explore how the results from a common strategy for handling missing post-treatment follow-up data [5-7] compared to those from the IP-weighting approach. In particular, we repeated the analysis described in the previous section, this time without applying IP weights, thereby imposing the assumption that people with and without FU are similar on average with regard to factors that influence recurrence. This approach does not account for the potential selection bias induced by excluding patients who lack FU data

#### Simulation Study

Finally, to investigate the impact of potential selection bias in a different data setting, we conducted a simulation study (see Supporting Information). Briefly, we generated an underlying population in which cavitary disease is strongly associated with having missing FU, TB recurrence, and death. We then computed and compared estimates of overall TB recurrence risk with and without adjustment for potential selection bias, i.e., with and without weights to account for joint predictors of missing FU and TB recurrence. We investigated the impact on results of varying different characteristics of the baseline simulation scenario, such as the underlying risk of TB recurrence.

## RESULTS

Of the 2,762 patients with confirmed RR/MDR-TB who initiated a longer individualized regimen containing BDQ and/or DLM, 2,053 (74.3%) patients were cured or completed treatment (Figure 2). After removing patients from North Korea, the final dataset we analyzed contained 1,991 patients. Among these individuals, 17 (0.9%) died within six-months post-treatment, 746 (37.5%) lacked post-treatment follow-up data, and 1228 (61.7%) had an observed six-month post treatment-outcome of no change or recurrence.

**Figure 2:**
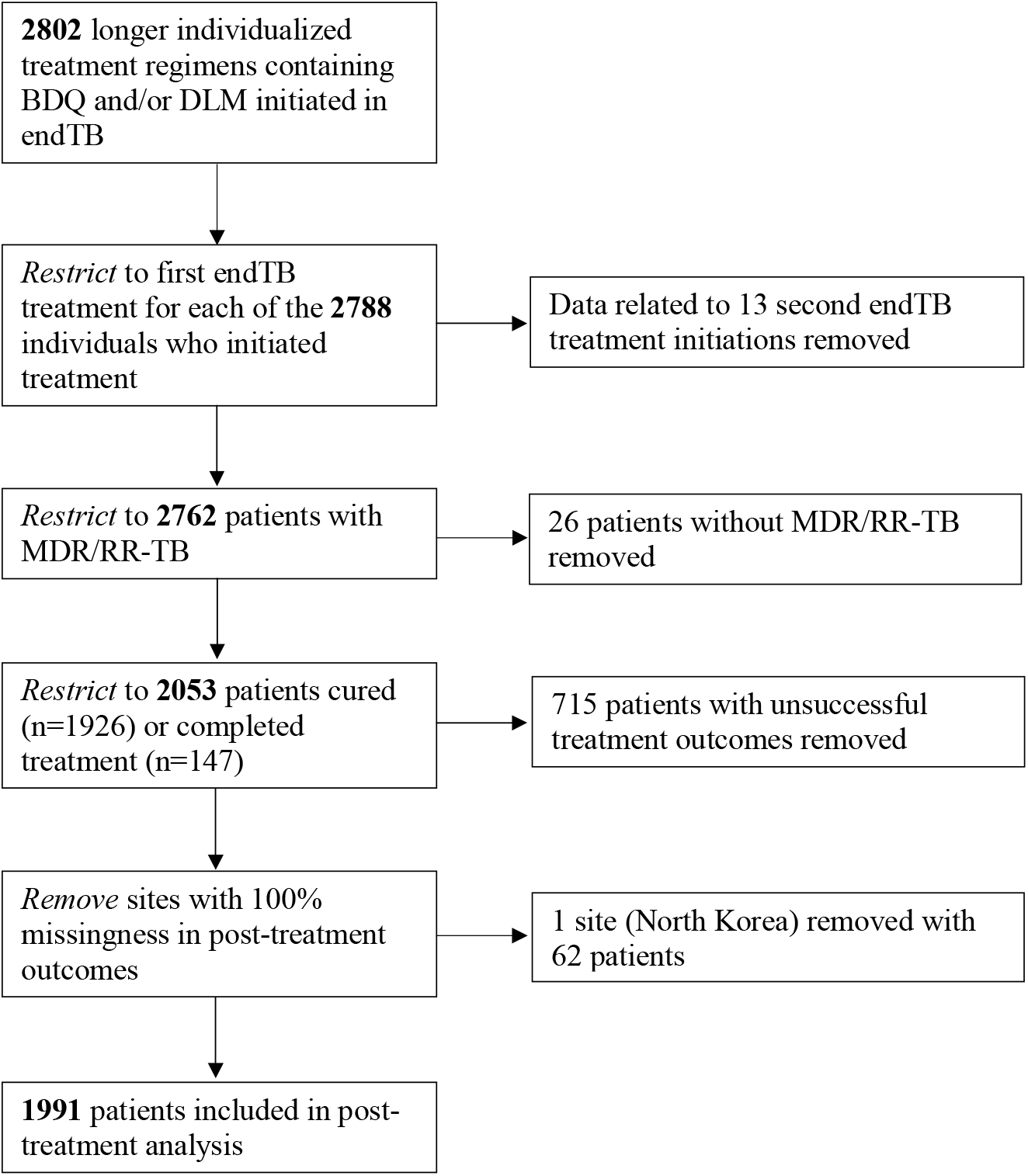
Inclusion/exclusion criteria for the post-treatment analysis

Table 2 shows the characteristics of the 1,991 patients, overall and by six-month post-treatment outcome. Overall, 11.9% of patients were living with HIV; 21.4% of those who lacked six-month follow-up data and 11.8% of those who died post-treatment had HIV coinfection. The overall patient age distribution was similar to that among patients who were missing FU data or were observed at six-months post-treatment, while patients who died were more likely to be aged 60 or older. Compared to patients with follow-up data, patients who lacked follow-up data were more likely to have a body mass index <18.5 and were less likely to have diabetes. The rate of post-treatment deaths and missing follow-up was variable across sites.

**Table 2:**
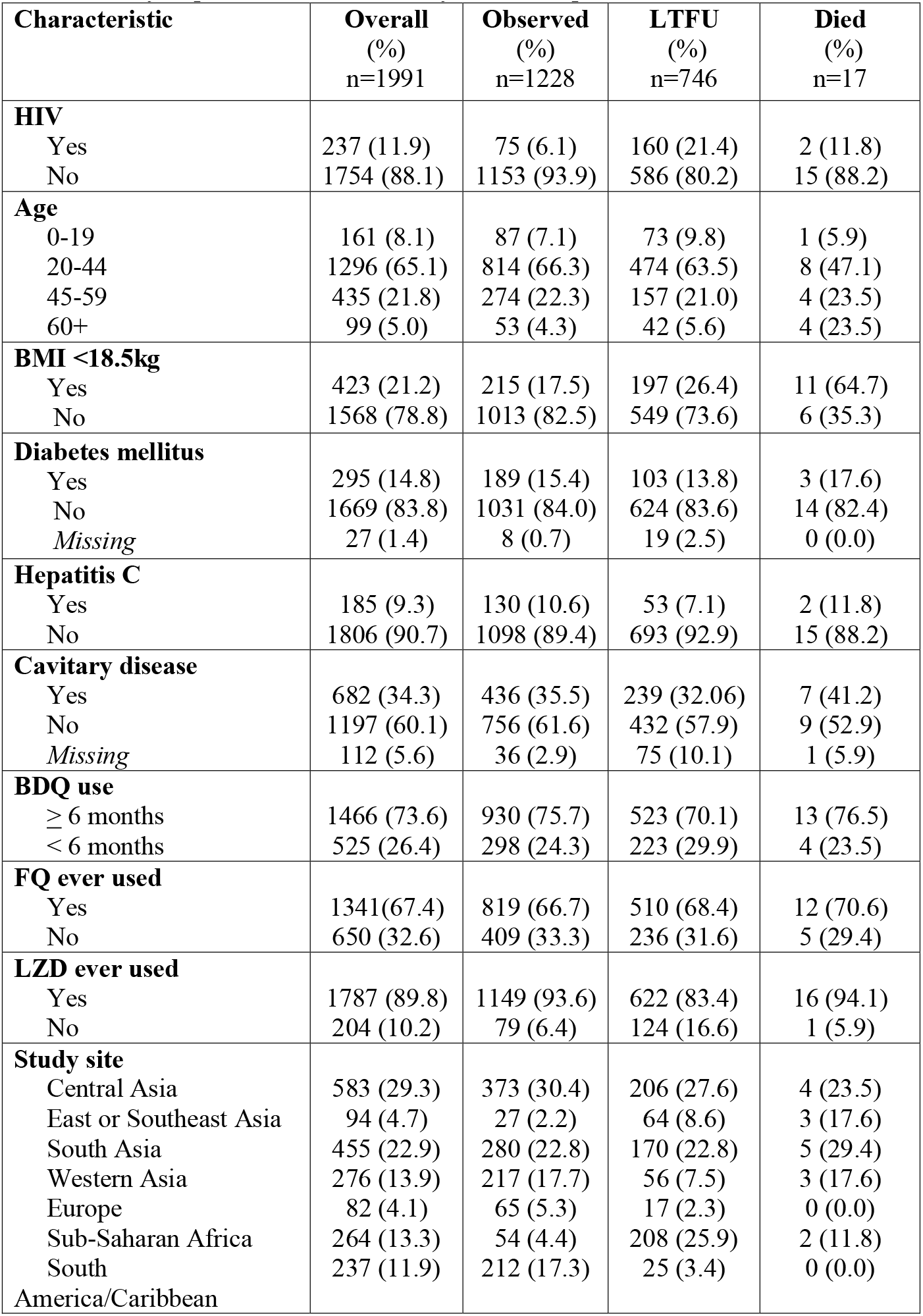
Summary of patient characteristics by six-month post-treatment outcome.

### Six-month risk of TB recurrence

Ten patients (0.5%) experienced recurrent TB within six-months post-treatment with a median time to recurrence of 161 days. Table 3 shows the estimated six-month risk of recurrent TB under the five strategies for handling post-treatment deaths. The risk of recurrent TB was 6.6 (95% credible interval [CI]: 3.2, 11.2) per 1000 individuals under the *observed recurrence* strategy, and 6.7 (95% CI: 2.8, 12.2) per 1000 under the *hypothetical recurrence* strategy. The estimated frequencies of a composite outcome under the *observed composite/any, observed composite/probable*, and *observed composite/explicit* strategies were 24.2 (95% CI: 14.1, 37.0), 10.5 (95% CI: 5.6, 16.6), and 7.8 (95% CI: 3.9, 13.2) per 1000, respectively.

**Table 3:** Estimated risk of recurrence overall and by HIV status under five strategies for handling post-treatment deaths.

| Strategy | Risk per 1000<sup>1</sup> |  |  | Relative risk<sup>1</sup> |
| --- | --- | --- | --- | --- |
|  | Overall | With HIV | Without HIV |  |
| **Observed recurrence** | 6.6<br>(3.2, 11.2) | 9.2<br>(2.0, 31.7) | 6.3<br>(2.9, 10.8) | 1.46<br>(0.31, 7.08) |
| **Hypothetical recurrence** | 6.7<br>(2.8, 12.2) | 9.0<br>(2.1, 33.5) | 6.4<br>(2.2, 11.5) | 1.42<br>(0.30, 7.14) |
| **Observed composite/any** | 24.2<br>(14.1, 37.0) | 15.6<br>(2.8, 42.4) | 25.3<br>(14.6, 40.0) | 0.62<br>(0.12, 2.01) |
| **Observed composite/probable** | 10.5<br>(5.6, 16.6) | 9.2<br>(2.1, 32.6) | 10.7<br>(5.1, 16.7) | 0.85<br>(0.20, 3.86) |
| **Observed composite/explicit** | 7.8<br>(3.9, 13.2) | 9.2<br>(2.1, 32.6) | 7.6<br>(3.4, 13.1) | 1.20<br>(0.25, 5.73) |

<sup>1</sup>95% credible intervals are constructed as the 2.5<sup>th</sup> and 97.5<sup>th</sup> percentiles of the 1000 bootstrap sample estimates

### Risk of TB recurrence by HIV status

The relative risk of recurrent TB in patients with and without HIV under the *observed recurrence* and *hypothetical recurrence* strategies were 1.46 (95% CI: 0.31, 7.08) and 1.42 (95% CI: 0.30, 7.14), respectively (Table 3). In both cases, relative risks were above one, suggesting a higher risk of recurrence among patients living with HIV; however, confidence intervals were wide. The corresponding risk ratios for the three composite outcomes were 0.62 (95% CI: 0.12, 2.01), 0.85 (95% CI: 0.20, 3.86), and 1.20 (95% CI: 0.25, 5.73) for the respectively *observed composite/any, observed composite/probable*, and *observed composite/explicit* strategies, respectively.

### Sensitivity analysis

When patients missing post-treatment follow-up were excluded from the analysis without adjustment for potential selection bias, the estimated risk of recurrent TB overall was 7.5 per 1000 (95% CI: 3.9, 11.2) under the *observed recurrence* strategy, 7.5 per (95% CI: 3.1, 11.8) under the *hypothetical recurrence* strategy and 20.2 (95% CI: 13.1, 29.3), 11.9 (95% CI: 6.6, 18.3), and 9.1 (95% CI: 4.8, 15.4) per 1000 under the three composite strategies (Table 4).

**Table 4:** Sensitivity analysis results; estimated risk of recurrence overall and by HIV status under five strategies for handling post-treatment deaths, without IP-weighting to account for excluded patients with missing FU data.

| Strategy | Risk per 1000<sup>1</sup> |  |  | Relative risk<sup>1</sup> |
| --- | --- | --- | --- | --- |
|  | Overall | With HIV | Without HIV |  |
| **Observed recurrence** | 7.5<br>(3.9, 11.2) | 14.0<br>(4.3, 44.3) | 6.6<br>(3.2, 11.0) | 2.13<br>(0.64, 9.12) |
| **Hypothetical recurrence** | 7.5<br>(3.1, 11.8) | 13.3<br>(4.2, 42.0) | 6.7<br>(2.4, 12.3) | 1.99<br>(0.56, 10.14) |
| **Observed composite/any** | 20.2<br>(13.1, 29.3) | 25.9<br>(5.9, 65.0) | 19.4<br>(11.8, 28.3) | 1.33<br>(0.31, 3.47) |
| **Observed composite/probable** | 11.9<br>(6.6, 18.3) | 14.0<br>(4.4, 42.2) | 11.7<br>(5.8, 17.8) | 1.20<br>(0.38, 4.38) |
| **Observed composite/explicit** | 9.1<br>(4.8, 15.4) | 14.0<br>(4.4, 42.2) | 8.4<br>(3.4, 14.3) | 1.66<br>(0.47, 6.63) |

<sup>1</sup>95% credible intervals are constructed as the 2.5<sup>th</sup> and 97.5<sup>th</sup> percentiles of the 1000 bootstrap sample estimates

The impact of potential selection bias was more apparent when looking at estimates of relative risk by HIV status. When results were not adjusted for selection bias, the point estimates differed from the adjusted estimates in terms of magnitude, and in some cases, direction (Table 4).

### Simulation study

The results of the simulation study are shown in Table S3-S9 of the Supporting Information. The variation in results across simulation scenarios demonstrates that there are several factors that increase the impact of selection bias, such as a stronger relationship between the joint predictor(s) of missing FU and recurrence and 1) missing FU or 2) recurrence.

## DISCUSSION

The overall risk of TB recurrence, or risk of TB recurrence or death, was low in this population, substantiating the effectiveness of longer MDR-TB regimens containing BDQ and/or DLM. This was true irrespective of method used to handle post-treatment deaths. The association between HIV and TB recurrence was inconclusive due to the small number of recurrences and deaths, which resulted in wide confidence intervals across all strategies.

The estimates of TB recurrence and relative risk comparing patients with and without HIV were similar whether derived under the assumption that patients who died post-treatment did not experience recurrent TB (*observed recurrence*) or in a hypothetical scenario where all patient deaths could have been prevented (*hypothetical recurrence*). There are other settings in which the difference between these two strategies might be more pronounced; for example, if post-treatment deaths were more common and patients who died had a much higher risk of recurrent TB compared to patients who did not, then the *hypothetical recurrence* estimate would be markedly higher than the *observed recurrence* estimate.

As expected, estimates of the six-month incidence of composite outcomes of TB recurrence or death were higher than the estimates for recurrence alone. Furthermore, estimates varied widely depending on which deaths were included in the composite outcome: the incidence of recurrent TB or death from any cause was twice that of recurrent TB or TB-related death, and the risk ratio estimate changed direction depending on whether deaths of unknown cause were included in the outcome or assumed to be non-recurrences. The proportion of post-treatment deaths that are related to TB, and therefore, the importance of incorporating cause of death into composite estimates, will vary by setting and highlights the importance of ascertaining and recording such information.

The estimates from each strategy answer different questions and in practice, researchers will need to select which one to report [1-4]. Intuitively, the *observed recurrence* strategy may be preferred to the *hypothetical recurrence* strategy, as the latter considers a scenario that is aspirational but not currently relevant to most settings (i.e., one in which all post-treatment deaths could be prevented). On the other hand, the *observed recurrence* strategy may be inadequate in certain circumstances. For example, if patients with a certain characteristic were much more likely to die post-treatment compared to those without that characteristic, the *observed recurrence* estimate could indicate that patients with the characteristics are less likely to experience recurrent TB just because earlier death largely prevented this group from developing recurrent TB; see Rojas-Saunero et. al [2] for another example of this. A second scenario in which the *observed recurrence* strategy may not be ideal is when deaths due to undetected TB are common. In this case, one of the composite strategies might be preferable, particularly when complete information on cause of death is available. One potential drawback of the composite strategies is that they change the outcome of interest, which may not always be of scientific interest.

Patients with low BMI or HIV were more likely, and patients with diabetes were less likely, to have missing FU data in this study. Previous studies have linked each of these subgroups to an increased risk of recurrent TB [8,10,17], which underscores the importance of accounting for patients with missing FU data when estimating recurrent TB. In our sensitivity analysis, the estimates that were computed by excluding patients with missing FU data but without accounting for selection bias through IP-weighting were similar to the IP-weighted estimates of overall recurrence. The differences in the weighted and unweighted relative risk estimates were more pronounced, though wide confidence intervals resulted in the same conclusion. This will not always be the case: larger imbalances in the characteristics of patients with and without FU combined with a stronger association of these characteristics with TB recurrence risk would result in a greater impact of selection bias, and consequently larger differences in the weighted and unweighted estimates. The simulation study of the Supporting Information provides examples of several such settings.

This study has several limitations. First, we were not able to distinguish between TB relapse and reinfection. While the rate of reinfection will vary by setting, we sought to limit misclassification by estimating the six-month incidence, a time during which relapse is likely to be the prevailing source of recurrent TB [28]. Second, we considered a less informative binary outcome rather than a time to event outcome [6], as follow-up was only consistently reported using a six-month post-treatment form at most sites rather than through continuous follow-up. Third, our results on HIV as a determinant of recurrence may be affected by collider bias [29-30]. This could occur if there is a factor such as age that is associated with both HIV status and inclusion in the cohort. If a third factor, such as TB disease severity, is associated with cohort inclusion and recurrent TB risk, the relationship observed between HIV status and recurrent TB may be distorted. This is a challenge with all observational TB studies, as TB is more likely to go undetected among people living with HIV [31]. Therefore, this group is less likely to be included in a treatment cohort.

This bias could be reduced through IP-weighting if data were available on factors related to 1) to HIV status and cohort inclusion and 2) cohort inclusion and experiencing post-treament TB recurrence. Unfortunately, this information is often unattainable in practice, as it requires being able to identify all people living with HIV who are eligible for MDR-TB treatment.

As multidrug- and rifampicin-resistant tuberculosis (MDR/RR-TB) treatment guidelines evolve in response to emerging evidence [32-34] on the relative safety and effectiveness of all-oral shortened regimens compared to conventional longer treatments, an important and currently unanswered question is whether the relative safety and effectiveness of shortened vs longer regimens differ across important patient subgroups, such as people living with HIV [35]. Rigorous analyses of post-TB treatment recurrence are needed to accurately understand whether certain groups may require closer monitoring, given possible increased risk of relapse [36-38] relative to longer regimens. While methods for handling competing events and potential selection bias due to losses to follow-up are well-established [4, 19-20], their use in the context of estimating TB recurrence is sparse. Careful handling of post-treatment deaths and missing post-treatment follow-up data together with accompanying sensitivity analyses will generate evidence that is more robust and interpretable, with the overall goal of improving treatment and care.

## Data Availability

Some of the data included in this analysis are managed in countries governed by the European Union General Data Protection Regulation (GDPR). The data contain sensitive and potentially identifying information and cannot be sufficiently anonymized to meet GDPR standards and retain their utility. Pseudo-anonymized data will be made available upon request to an MSF Medical Director at, and execution of a data sharing agreement or alternate means that allows assurance that principles of GDPR regulations will be met.

## Acknowledgements

The authors thank the patients who participated in the endTB Observational Study and the clinicians and program staff of participating national tuberculosis programs. They also thank the endTB staff at Partners in Health, Doctors Without Borders, Epicentre, and Interactive Research and Development.

## Author contributions

SMS, CDM, and MF led the conceptualization, data curation, methodology, analysis, and writing of the paper. CDM, MF, MR, KS, UK, PK MB, CH, and HH led the design of the endTB study. UK, CH, MB, DH, SL, MK, SP, SA, AKI, A Krisnanda, SCV, SB, A Kumsa, WD, KT, MM, HA, TV, TTT, KS, MR, HH, and PK contributed to data collection and curation. All authors contributed to editing and review of the paper.

